# GLP-1 Receptor Agonists Added to Statin Therapy and Cardiovascular Outcomes in ASCVD: A Propensity Score–Matched Cohort Study

**DOI:** 10.64898/2026.11.05.26367396

**Authors:** Sunanda Tah, Michael Cheshire

**Affiliations:** Department of Internal Medicine, West Virginia School of Medicine – Camden Clark Medical Center

## Abstract

**Background:** Adding a GLP-1 receptor agonist (GLP-1 RA) to statin therapy in patients with atherosclerotic cardiovascular disease (ASCVD) and obesity may reduce cardiovascular risk, yet real-world evidence is limited.

**Objectives:** To evaluate whether GLP-1 RA use added to statins is associated with lower all-cause mortality, myocardial infarction (MI), and ischemic stroke versus statin monotherapy in ASCVD patients with body mass index (BMI) ≥30 kg/m^2^.

**Methods:** Using the TriNetX Research Network (108 U.S. healthcare organizations), we identified ASCVD patients with BMI ≥30 kg/m^2^ on statins initiating a GLP-1 RA (n=166,278) versus statin-only controls (n=920,176). Propensity score matching on 34 covariates yielded 119,911 matched pairs followed ≤3 years. Prespecified analyses included a BMI ≥35 subgroup (62,385 pairs), four washout sensitivity analyses (30–120 days), and a familial hypercholesterolemia exclusion analysis (161,312 matched pairs).

**Results:** GLP-1 RA use was associated with significantly lower all-cause mortality (4.0% vs. 7.5%; HR 0.479; 95% CI: 0.463–0.496; p<0.001), MI (HR 0.762; 95% CI: 0.737–0.788; p<0.001), and ischemic stroke (HR 0.671; 95% CI: 0.588–0.766; p<0.001). Benefits were consistent across the BMI ≥35 subgroup and all washout analyses. GLP-1 RA users were more likely to achieve LDL <55 mg/dL by risk analysis (17.1% vs. 15.3%; RR 1.118; p<0.001); the Kaplan-Meier analysis was non-significant (HR 1.007; p=0.538). Mortality benefit was preserved in the FH exclusion analysis.

**Conclusions:** ASCVD patients with obesity on statins, GLP-1 RA use was associated with substantially lower mortality, MI, and stroke, suggesting GLP-1 RAs may serve as cardioprotective adjuncts to statin therapy in this population.

**Graphical Abstract:** 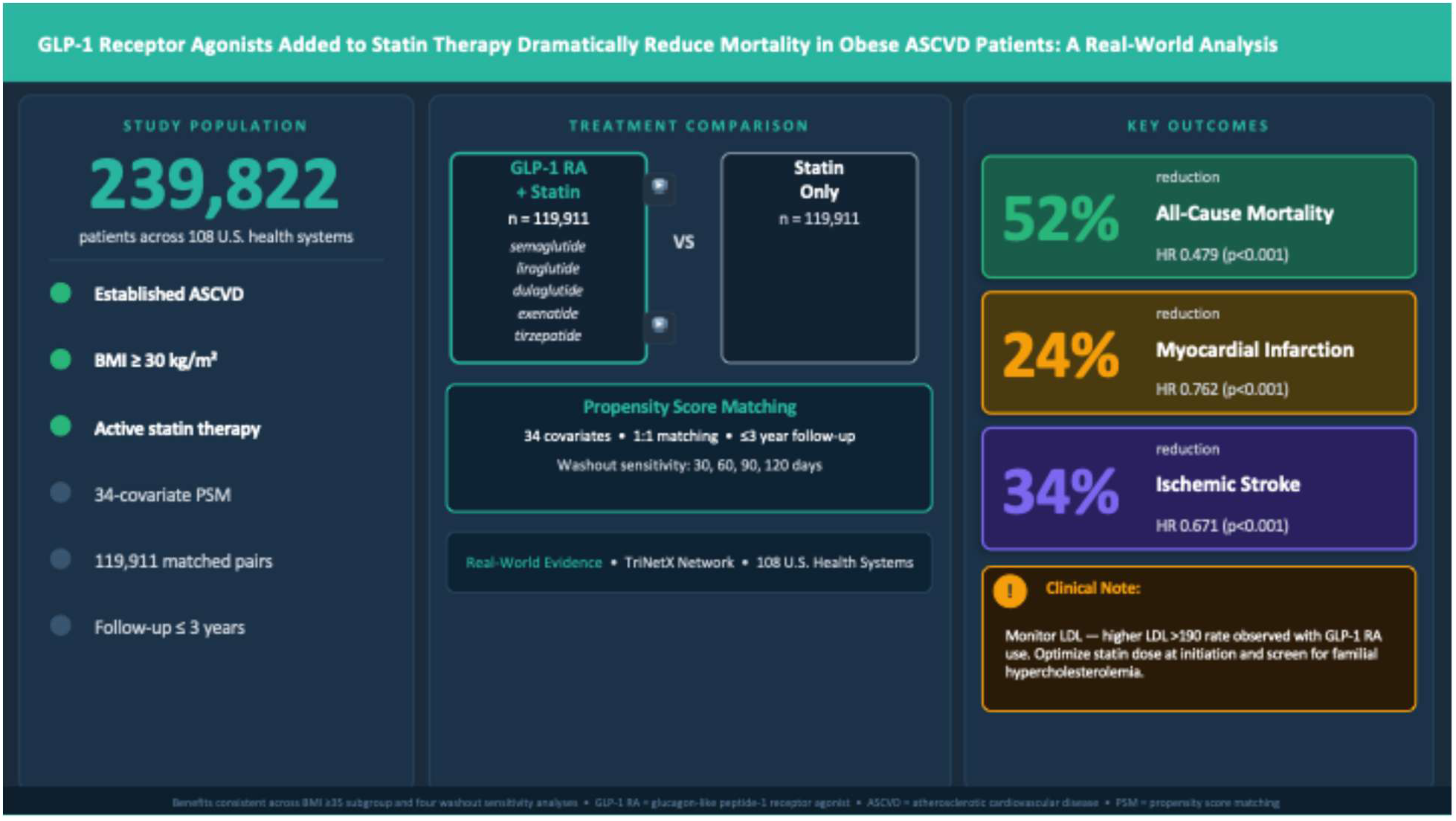

## INTRODUCTION

Atherosclerotic cardiovascular disease (ASCVD) remains the leading cause of death worldwide.^1^ Statins have revolutionized cardiovascular risk reduction, yet substantial residual risk persists even at guideline-recommended LDL targets.^2^ This gap is particularly wide in patients with obesity, which affects more than 40% of U.S. adults^3^ and independently fuels chronic inflammation, insulin resistance, and atherogenic dyslipidemia.^4^

GLP-1 receptor agonists (GLP-1 RAs) semaglutide, liraglutide, dulaglutide, exenatide, and tirzepatide (a dual GIP/GLP-1 receptor agonist) were developed for glycemic management but demonstrated cardiovascular benefit in landmark outcome trials. LEADER,5 SUSTAIN-6,6 and REWIND7 each showed MACE reduction in high-risk diabetic populations. Critically, SELECT enrolled non-diabetic patients that were overweight or obese, and had established ASCVD, demonstrated a 20% MACE reduction with semaglutide 2.4 mg, establishing cardiovascular benefit independent of glycemia.8

Despite these trial data, large-scale real-world evidence characterizing the incremental benefit of adding GLP-1 RAs to guideline-directed statin therapy is lacking. Most ASCVD patients are already on statins in clinical practice, and whether combination therapy reduces mortality, MI, and stroke at the population level has not been rigorously characterized.^9,10^

Therefore, we conducted a large, propensity score matched retrospective cohort study using the TriNetX federated research network to evaluate the association of GLP-1 RA therapy added to statins versus statin monotherapy on cardiovascular outcomes in ASCVD patients with elevated BMI.

## METHODS

### Data Source

We used the TriNetX Research Network, a federated platform providing de-identified electronic health record (EHR) data from 108 U.S. healthcare organizations encompassing diagnoses (ICD-10-CM), medications (RxNorm), procedures (ICD-10-PCS), laboratory results, and demographics. This study used aggregate de-identified data; institutional review board approval was not required under federal regulations governing de-identified data.

### Study Population

Cohort 1 (GLP-1 RA + statin) comprised patients with ASCVD, BMI ≥30 kg/m^2^, and concurrent statin and GLP-1 RA use. Cohort 2 (statin only) comprised patients with ASCVD, BMI ≥30 kg/m^2^, statin use, and no GLP-1 RA record. ASCVD was defined by at least one ICD-10-CM code: old MI (I25.2), atherosclerotic heart disease of native coronary artery (I25.1), ischemic cardiomyopathy (I25.5), aortocoronary bypass graft status (Z95.1), or coronary angioplasty with implant (Z95.5). Statins included atorvastatin, rosuvastatin, simvastatin, pravastatin, lovastatin, fluvastatin, and pitavastatin. GLP-1 RAs included semaglutide, liraglutide, dulaglutide, exenatide, and tirzepatide (a dual GIP/GLP-1 receptor agonist). All cohorts excluded patients with type 1 diabetes (E10), prior bariatric surgery (Z98.84), or pregnancy (ICD-10-PCS group 10).

### Index Event and Follow-up

The index event for Cohort 1 was the first recorded GLP-1 RA prescription; for Cohort 2, the first statin prescription without any GLP-1 RA record. Outcomes were captured from 1 day to 1,095 days post-index. Patients were censored after the last recorded EHR fact. Events prior to the time window were excluded from each outcome analysis.

### Propensity Score Matching

Propensity score matching (PSM) was performed across 34 pre-specified covariates: current age, age at index, sex, race (White, Black, American Indian/Alaska Native, Native Hawaiian/Pacific Islander, Asian, Other, Unknown), ethnicity (Hispanic, Non-Hispanic, Unknown), essential hypertension, type 2 diabetes, heart failure, chronic kidney disease (stages 3–5), peripheral vascular disease, ischemic stroke history, transient ischemic attack, tobacco use, hyperlipidemia, atrial fibrillation, long-term antiplatelet use, homelessness, prior inpatient encounters, and concurrent beta-blockers, ACE inhibitors, metformin, insulin, SGLT2 inhibitors, ezetimibe, evolocumab, alirocumab, inclisiran, bempedoic acid, aspirin, and P2Y12 inhibitors (clopidogrel, ticagrelor). Balance was assessed by standardized mean differences (SMDs); SMD <0.10 indicated adequate balance.

### Outcomes

Co-primary outcomes were: (1) all-cause mortality, (2) acute MI (ICD-10-CM I21, I21.3), and (3) ischemic stroke (I63.50), each excluding patients with the outcome prior to the time window. Secondary outcomes were: (4) incident LDL >190 mg/dL and (5) incident LDL <55 mg/dL, both defined using most-recent laboratory values within the time window. Because LDL represents a cross-sectional laboratory measurement rather than a traditional time-to-event endpoint, risk analysis (proportion of eligible patients meeting the threshold during follow-up) was pre-specified as the primary analysis for these outcomes. Kaplan-Meier survival analysis is reported as a secondary analysis and reflects time to first recorded qualifying LDL value, which is subject to influence by laboratory testing frequency. Denominators for LDL analyses differ from the primary matched cohort as patients with the respective LDL threshold met prior to the index event are excluded per TriNetX protocol.

### Statistical Analysis

For each outcome, we report risks (proportions), risk difference, risk ratio (RR) with 95% CIs and two-sided z-test p values, and odds ratio. Kaplan-Meier survival analysis with log-rank testing was performed. Cox proportional hazards regression yielded hazard ratios (HRs) with 95% CIs; proportionality was assessed via Schoenfeld residuals. A prespecified subgroup analysis restricted to BMI ≥35 kg/m^2^ (62,385 matched pairs) was performed. Four sensitivity analyses varied the washout window to 30, 60, 90, and 120 days post-index. All analyses were conducted on propensity-matched cohorts. P values are reported to three decimal places; p<0.05 was the threshold for statistical significance. Patients with missing outcome data were excluded from the respective analysis, with exclusion counts reported. Analyses were performed using the TriNetX Compare Outcomes platform. A prespecified sensitivity analysis excluding patients with a most-recent LDL ≥190 mg/dL from both cohorts prior to the index event was performed as a surrogate exclusion for familial hypercholesterolemia (FH exclusion analysis), to assess robustness of lipid and cardiovascular outcomes in a population without extreme baseline LDL elevation.

## RESULTS

### Study Cohorts and Propensity Score Matching

Before matching, Cohort 1 comprised 166,278 patients (GLP-1 RA + statin, BMI ≥30) and Cohort 2 comprised 920,176 patients (statin only, BMI ≥30). Pre-match, Cohort 1 was younger (mean age 62.6 vs. 64.5 years, SMD 0.174), more often female (44.2% vs. 38.7%), and had substantially higher rates of type 2 diabetes (64.0% vs. 22.8%), hypertension (75.9% vs. 47.5%), and use of glucose-lowering agents—reflecting the indication bias intrinsic to GLP-1 RA prescribing patterns.

After 1:1 PSM, 119,911 matched pairs were retained. All 34 covariates achieved SMD <0.10 post-match (Table 1). Median follow-up was 895 days for the GLP-1 RA group and 739 days for the statin-only group.

**Table 1.** Baseline characteristics of propensity score–matched cohorts (BMI ≥30, N=119,911 per group).

| **Characteristic** | **GLP-1 RA + Statin**<br>**(N=119,911)** | **Statin Only**<br>**(N=119,911)** | **SMD** |
|---|---|---|---|
| **Demographics** |  |  |  |
| Age at index, mean ± SD, y | 63.0 ± 10.1 | 62.8 ± 11.6 | 0.022 |
| Female sex, n (%) | 53,039 (44.2%) | 53,567 (44.7%) | 0.009 |
| White race, n (%) | 88,853 (74.1%) | 89,257 (74.4%) | 0.008 |
| Black race, n (%) | 16,944 (14.1%) | 16,638 (13.9%) | 0.007 |
| **Comorbidities** |  |  |  |
| Type 2 diabetes mellitus, n (%) | 62,426 (52.1%) | 63,694 (53.1%) | 0.021 |
| Essential hypertension, n (%) | 83,264 (69.4%) | 80,902 (67.5%) | 0.042 |
| Heart failure, n (%) | 24,272 (20.2%) | 22,764 (19.0%) | 0.032 |
| Chronic kidney disease, stage 3, n (%) | 12,713 (10.6%) | 12,092 (10.1%) | 0.017 |
| Atrial fibrillation, n (%) | 13,755 (11.5%) | 12,889 (10.7%) | 0.023 |
| **Concurrent medications** |  |  |  |
| Metformin, n (%) | 25,596 (21.3%) | 25,550 (21.3%) | 0.001 |
| Insulin, n (%) | 32,552 (27.1%) | 32,086 (26.8%) | 0.009 |
| SGLT2 inhibitor, n (%) | 8,304 (6.9%) | 7,658 (6.4%) | 0.022 |
| Beta-blocker, n (%) | 54,158 (45.2%) | 52,569 (43.8%) | 0.027 |
| Ezetimibe, n (%) | 4,876 (4.1%) | 4,469 (3.7%) | 0.018 |
| **Follow-up** |  |  |  |
| Median follow-up, days (IQR) | 895 (716) | 739 (844) | — |

*CKD = chronic kidney disease; IQR = interquartile range; PSM = propensity score matching; SGLT2 = sodium-glucose cotransporter-2; SMD = standardized mean difference. All SMDs <0.10 indicate adequate post-match balance.*

### Primary Outcomes

Results are presented in Table 2. GLP-1 RA use was associated with significantly lower all-cause mortality (4.0% vs. 7.5%; RR 0.527 (95% CI: 0.509–0.545); HR 0.479 (95% CI: 0.463–0.496); p<0.001), corresponding to a 52% relative mortality reduction. Three-year survival probability was 94.2% vs. 89.2% (log-rank χ^2^=1,755.2, p<0.001).

**Table 2.** Primary outcomes: GLP-1 RA + statin versus statin only (BMI ≥30, N=119,911 matched pairs).

| **Outcome** | **GLP-1 RA + Statin Events/Risk** | **Statin Only Events/Risk** | **Risk Ratio (95% CI)** | **Hazard Ratio (95% CI)** | **P Value** |
|---|---|---|---|---|---|
| **All-cause mortality** | 4,733 / 4.0% | 8,986 / 7.5% | 0.527 (0.509–0.545) | 0.479 (0.463–0.496) | < 0.001 |
| **Myocardial infarction** | 6,241 / 6.0% | 7,155 / 7.1% | 0.843 (0.816–0.871) | 0.762 (0.737–0.788) | < 0.001 |
| **Ischemic stroke** | 377 / 0.3% | 520 / 0.4% | 0.730 (0.640–0.833) | 0.671 (0.588–0.766) | < 0.001 |
| **LDL >190 mg/dL** | 1,069 / 0.9% | 909 / 0.8% | 1.196 (1.095–1.306) | 1.084 (0.992–1.184) | 0.074† |
| **LDL <55 mg/dL** | 15,979 / 17.1% | 16,557 / 15.3% | 1.118 (1.096–1.141) | 1.007 (0.985–1.029) | 0.538‡ |

*CI = confidence interval; HR = hazard ratio; LDL = low-density lipoprotein; RR = risk ratio. P values two-tailed. LDL outcome denominators differ from the primary matched cohort (N=119,911) because patients with the respective LDL outcome prior to the index event are excluded per TriNetX protocol (LDL <55: 93,737 eligible in C1, 108,220 in C2; LDL >190: 118,842 eligible in C1, 119,002 in C2). Risk analysis is the primary analysis for LDL outcomes given the snapshot nature of laboratory values; survival analysis reflects time to first recorded LDL meeting threshold and is influenced by testing frequency. LDL <55 HR non-significant (p=0.538); LDL >190 HR non-significant (p=0.074).*

MI risk was significantly lower in GLP-1 RA users (6.0% vs. 7.1%; RR 0.843 (95% CI: 0.816– 0.871); HR 0.762 (95% CI: 0.737–0.788); p<0.001). Ischemic stroke likewise occurred less frequently (0.3% vs. 0.4%; RR 0.730 (95% CI: 0.640–0.833); HR 0.671 (95% CI: 0.588–0.766); p<0.001).

### Lipid Outcomes

LDL outcome analyses were performed on patients without the respective LDL outcome prior to the index event, resulting in reduced denominators that differ from the primary matched cohort (LDL <55 analysis: 93,737 eligible in Cohort 1 vs. 108,220 in Cohort 2; LDL >190 analysis: 118,842 vs. 119,002). Risk analysis was pre-specified as the primary analysis for LDL outcomes, given that laboratory values represent cross-sectional measurements rather than traditional time-to-event endpoints; the Kaplan-Meier analysis reflects time to first recorded LDL meeting the threshold and is subject to testing frequency bias. GLP-1 RA users were significantly more likely to achieve the guideline-recommended LDL <55 mg/dL target by risk analysis (17.1% vs. 15.3%; RR 1.118; 95% CI: 1.096–1.141; p<0.001). The survival analysis was non-significant (HR 1.007; 95% CI: 0.985–1.029; p=0.538), indicating that while more GLP-1 RA users achieved this threshold at any point during follow-up, the timing of LDL testing did not differ substantially between groups. This finding is consistent with GLP-1 RA therapy facilitating guideline lipid target attainment through weight-mediated improvements in lipid homeostasis and more intensive cardiometabolic monitoring at initiation. A modest increase in incident LDL >190 mg/dL was observed by risk analysis in GLP-1 RA users (0.9% vs. 0.8%; RR 1.196; 95% CI: 1.095–1.306; p<0.001). This was not confirmed by survival analysis (HR 1.084; 95% CI: 0.992–1.184; p=0.074). Given the discordance between analyses, this finding should be interpreted as a signal warranting clinical vigilance — particularly regarding statin optimization at GLP-1 RA initiation — rather than a confirmed causal outcome. Notably, this signal persisted in the FH exclusion sensitivity analysis (RR 1.690; p<0.001), confirming it is not entirely attributable to pre-existing familial hypercholesterolemia, though the altered denominator in that analysis limits direct comparability.

### BMI ≥35 kg/m^2^ Subgroup Analysis

In the prespecified subgroup of 62,385 matched pairs with BMI ≥35 kg/m^2^, findings were directionally consistent with the primary analysis. GLP-1 RA use remained associated with lower all-cause mortality (4.2% vs. 7.8%; HR 0.481 (95% CI: 0.459–0.505); p<0.001), MI (HR 0.797 (95% CI: 0.760–0.835); p<0.001), and ischemic stroke (HR 0.655 (95% CI: 0.546–0.785); p<0.001) (Table 3).

**Table 3.**
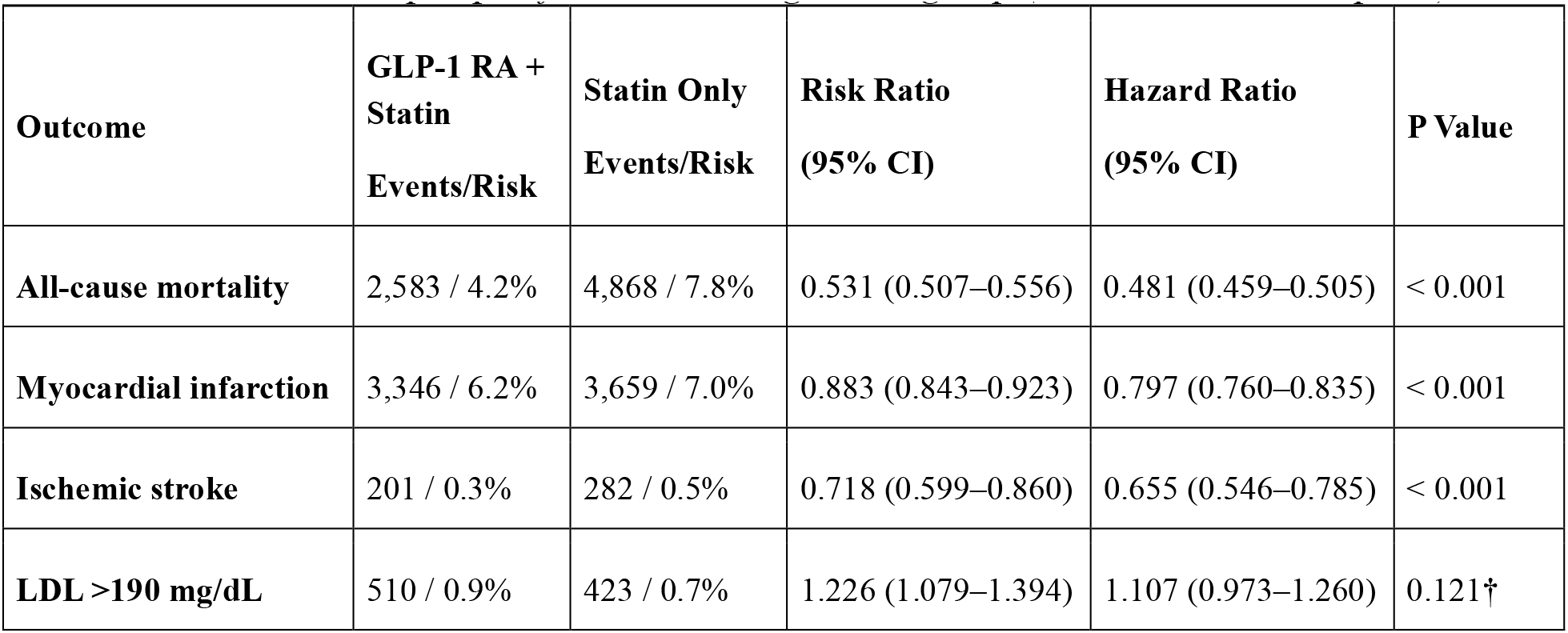
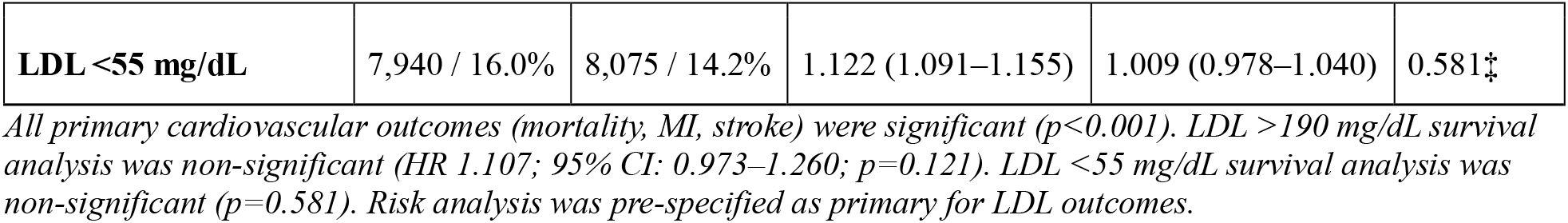
Outcomes in the prespecified BMI ≥35 kg/m^2^ subgroup (N=62,385 matched pairs).

### Sensitivity Analyses

Across washout windows of 30–120 days, GLP-1 RA use remained associated with significantly lower all-cause mortality (HR range: 0.555–0.611; all p<0.001) (Table 4). MI risk reduction weakened with longer washout periods but hazard ratios remained below 1.0 at every threshold, indicating durable cardioprotection. Stroke reduction was significant in the primary analysis; point estimates remained below 1.0 across all washout windows, though statistical significance was not maintained at 60 and 120 days. Familial Hypercholesterolemia Exclusion Sensitivity Analysis To address the potential confounding influence of familial hypercholesterolemia (FH) on LDL outcomes, a sensitivity analysis was performed excluding patients with a most-recent LDL ≥190 mg/dL from both cohorts prior to the index event (161,312 matched pairs). Mortality benefit was preserved and strengthened (3.2% vs. 7.9%; HR 0.348; 95% CI: 0.337–0.360; p<0.001). Ischemic stroke reduction was maintained (RR 0.897; HR 0.793; 95% CI: 0.712– 0.882; p<0.001). The LDL <55 mg/dL finding was substantially amplified (21.8% vs. 13.2%; RR 1.650; 95% CI: 1.622–1.678; p<0.001), further supporting the role of GLP-1 RA therapy in facilitating guideline lipid target achievement. MI risk reduction was attenuated in this analysis (HR 0.978; p=0.173), consistent with expected heterogeneity of treatment effect in a cohort pre-selected for lower baseline lipid burden. The LDL >190 mg/dL outcome in this analysis reflects incident LDL elevation above 190 mg/dL after baseline exclusion and should be interpreted with caution given the altered risk denominator.

**Table 4.** Sensitivity analyses: Mortality, MI, and stroke hazard ratios by washout window (BMI ≥30 cohort).

| Analysis | 1-Day (Primary) | 30-Day Washout | 60-Day Washout | 90-Day Washout | 120-Day Washout |
|---|---|---|---|---|---|
| **Matched pairs, n** | 119,911 | 118,318 | 118,303 | 115,034 | 118,318 |
| **Mortality HR (95% CI)** | 0.479 (0.463–0.496) | 0.555 (0.534–0.576) | 0.582 (0.559–0.605) | 0.592 (0.569–0.617) | 0.611 (0.586–0.636) |
| **MI HR (95% CI)** | 0.762 (0.737–0.788) | 0.850 (0.820–0.882) | 0.887 (0.854–0.921) | 0.902 (0.867–0.939) | 0.896 (0.861–0.933) |
| **Stroke HR (95% CI)** | 0.671 (0.588–0.766) | 0.788 (0.682–0.911) | 0.800 (0.687–0.932) | 0.827 (0.702–0.974) | 0.801 (0.681–0.942) |

*All HRs for GLP-1 RA + statin vs. statin only after PSM. Mortality HRs statistically significant (p<0.001) across all washout periods. HR = hazard ratio; MI = myocardial infarction; PSM = propensity score matching.*

## DISCUSSION

In this large propensity score–matched analysis of over 239,000 ASCVD patients with elevated BMI from 108 U.S. healthcare organizations, adding GLP-1 RA therapy to background statins was associated with a 52% relative reduction in all-cause mortality, 24% reduction in MI, and 34% reduction in ischemic stroke. The mortality signal was the most striking and most robust finding, consistent across BMI ≥35 subgroup and across all four washout sensitivity analyses, a pattern that strongly supports a true association rather than artifact.

Our findings extend prior cardiovascular outcome trial evidence to a real-world statin-treated ASCVD population. The LEADER,^5^ SUSTAIN-6,^6^ REWIND,^7^ and SELECT^8^ trials established GLP-1 RA cardiovascular benefit in controlled settings, but none specifically characterized benefit added to background statin therapy in a population-level electronic health record analysis. Our observed 52% relative mortality reduction substantially exceeds the 15–20% reductions seen in dedicated cardiovascular outcome trials.^9^ This likely reflects the broader ASCVD population captured by real-world data, the longer natural history of ASCVD, and the additive benefit of GLP-1 RAs on non-lipid cardiovascular risk factors incompletely addressed by statins.

Multiple mechanisms likely underlie these findings. Like statins, GLP-1 RAs reduce inflammation, but GLP-1 RAs have shown multiple other benefits, including positive effects on body weight, blood pressure, and, platelet aggregation.^12, 13^ Additionally, direct GLP-1 receptor signaling in cardiomyocytes and vascular endothelium attenuates ischemia-reperfusion injury and improves endothelial function.^14^ These complementary mechanisms, operating through pathways entirely distinct from LDL lowering, provide a compelling biological rationale for additive benefit in combination therapy.

A key secondary finding of this study is that GLP-1 RA users were significantly more likely to achieve the guideline-recommended LDL-C threshold of <55 mg/dL by risk analysis (17.1% vs. 15.3%; RR 1.118; 95% CI: 1.096–1.141; p<0.001). The Kaplan-Meier analysis was non-significant (HR 1.007; p=0.538), reflecting similar timing of LDL testing between groups rather than absence of a biologically meaningful difference. Risk analysis is the methodologically appropriate primary analysis for laboratory thresholds in federated EHR data, where survival curves reflect documentation frequency as much as clinical biology. This finding carries direct clinical importance. The 2026 ACC/AHA Dyslipidemia Guideline recommends an LDL-C goal of <55 mg/dL for very high-risk ASCVD patients on maximally tolerated statin therapy, a threshold associated with meaningful incremental reductions in MACE beyond what is achieved at higher LDL levels.10 The greater rate of LDL <55 mg/dL achievement in GLP-1 RA users likely reflects several mechanisms: GLP-1 RAs reduce hepatic lipogenesis and improve bile acid metabolism,15 weight loss of 5–15% reduces fasting triglycerides and can modestly lower LDL-C through improved hepatic cholesterol homeostasis, and the concurrent intensification of cardiometabolic monitoring that often accompanies GLP-1 RA initiation may prompt more aggressive statin dose titration. The absence of a significant time-to-event benefit on Kaplan-Meier analysis (HR 1.007; p=0.538), despite the significant risk analysis finding, suggests that GLP-1 RA users are more likely to have LDL <55 mg/dL at any point in follow-up, but that the timing of LDL target achievement does not differ substantially between groups. Clinically, this means GLP-1 RA therapy should be viewed not merely as an adjunct to statins for cardiovascular risk reduction, but as an agent that may actively facilitate attainment of the most stringent guideline lipid goals.

The LDL >190 mg/dL signal warrants careful interpretation. By risk analysis, GLP-1 RA users had a modestly higher incidence of incident LDL >190 mg/dL (RR 1.196; p<0.001), but this was not confirmed by survival analysis (HR 1.084; p=0.074). The discordance between these two analyses means this finding cannot be considered a confirmed outcome and should not be over-interpreted. It is best understood as a signal for clinical vigilance. Several mechanisms may contribute. Patients initiated on GLP-1 RAs could possibly also have familial hypercholesterolemia, as individuals with refractory dyslipidemia and complex cardiometabolic risk profiles are disproportionately referred for advanced therapies. GLP-1 RAs also exert pleiotropic effects on lipid metabolism, including alterations in hepatic cholesterol synthesis and bile acid signaling, that may differentially affect LDL-C in high risk individuals.15 Additionally, rapid weight loss induced by GLP-1 RAs may transiently mobilize adipose-stored cholesterol into the circulation, a phenomenon well-documented during major weight loss,16 before steady-state lipid remodeling occurs. At the clinical level, GLP-1 RA initiation may shift prescriber attention toward metabolic monitoring such as glycemic control, weight, and renal function, at the expense of statin dose titration. Notably, the higher LDL >190 mg/dL rate among GLP-1 RA users did not diminish the mortality benefit, suggesting that the broader cardiovascular effects of GLP-1 RAs including reduced inflammation, improved endothelial function, and direct heart muscle protection outweigh any added risk from elevated LDL in this population. Nonetheless, clinicians should maintain rigorous statin optimization concurrent with GLP-1 RA initiation, and patients with LDL >190 mg/dL should be evaluated for familial hypercholesterolemia and treated appropriately.10,17

The FH exclusion sensitivity analysis further supports the robustness of the primary findings. Excluding patients with baseline LDL ≥190 mg/dL — a well-validated surrogate for familial hypercholesterolemia in real-world data given the low sensitivity of ICD-10 coding for FH — preserved and amplified the mortality benefit (HR 0.348), reinforced the LDL <55 target achievement finding (RR 1.650), and maintained stroke reduction. The attenuation of MI benefit in this subgroup (HR 0.978; p=0.173) likely reflects the removal of a high-LDL subgroup in whom aggressive statin therapy already partially equalizes coronary risk between groups. Taken together, these findings suggest the cardiovascular benefits of GLP-1 RA therapy added to statins are not driven by a high-LDL minority and are robust to FH exclusion. The attenuation of the mortality HR from 0.479 (1-day washout) to 0.611 (120-day washout) across sensitivity analyses is informative. This gradient likely reflects an immortal time effect: patients who survive long enough to pass longer washout thresholds are selectively healthier at baseline.17 Even at the most stringent 120-day washout threshold, the mortality benefit remained large and statistically significant, suggesting a genuine effect rather than a methodological artifact.

In most GLP-1 RA cardiovascular outcome trials, stroke reduction has been modest relative to MI reduction. Notably, SUSTAIN-6 reported an unusual pattern in which ischemic stroke reduction (HR 0.61; 95% CI: 0.38–0.99) numerically exceeded the MI reduction (HR 0.74; 95% CI: 0.58–0.95),6 making it an outlier among the GLP-1 RA outcome trials. SELECT, by contrast, demonstrated a significant 28% reduction in non-fatal MI (HR 0.72; 95% CI: 0.61–0.85) alongside a non-significant trend toward stroke reduction, with coronary events driving the primary endpoint.8 In the present study, the observed MI reduction (HR 0.762) and stroke reduction (HR 0.671) were both statistically significant, and is consistent with the broader GLP-1 RA literature. The stroke signal attenuated more than the MI signal across longer washout sensitivity analyses, further suggesting that coronary risk modification is the more durable and central mechanism of GLP-1 RA cardiovascular protection in this ASCVD-enriched cohort. The current data should not be interpreted as evidence that stroke reduction is the primary driver of benefit, instead it is the cumulative reduction in coronary atherosclerotic events that accounts for the majority of the mortality benefit observed here.

### Study Limitations

Several limitations should be noted. First, residual confounding cannot be excluded despite PSM across 34 covariates; unmeasured factors including medication adherence, healthcare engagement, diet, exercise, and social determinants may contribute to observed associations. Second, the TriNetX platform provides aggregate de-identified data, precluding analysis of GLP-1 RA doses, statin intensity, and longitudinal laboratory trends. FH was approximated using LDL ≥190 mg/dL as a surrogate rather than formal ICD-10 coding (E78.01), as FH is substantially underdiagnosed in administrative data; the FH exclusion sensitivity analysis should be interpreted accordingly. Third, cause-specific mortality was unavailable. Fourth, results represent predominantly U.S. institutional populations and may not be generalizable to other settings. This study is hypothesis-generating; design limitations preclude causal inference.

## CONCLUSIONS

This analysis provides the largest real-world evidence to date that adding GLP-1 receptor agonists to statin therapy is associated with substantially lower mortality in obese ASCVD patients, a finding that warrants further investigation and may prompt a reassessment of how we treat this population. A 52% relative reduction in all-cause mortality, consistent across subgroups and four sensitivity analyses in over 239,000 matched patients, represents a clinically meaningful signal in this observational cohort. With residual cardiovascular risk persisting despite statin therapy and obesity endemic in the ASCVD population, GLP-1 RAs represent an increasingly important adjunct to guideline-directed statin therapy in this population. Clinicians may consider these findings when evaluating initiation of GLP-1 RAs in eligible obese ASCVD patients, maintain rigorous statin optimization, and monitor LDL given the paradoxical LDL >190 mg/dL signal. Prospective randomized trials to formally establish this combination as a standard cardiometabolic regimen are urgently needed.

## PERSPECTIVES

### What Is Known?

Statins significantly reduce cardiovascular events in ASCVD but leave substantial residual risk, amplified by obesity. GLP-1 receptor agonists reduce MACE in randomized trials, primarily in diabetic populations or patients with obesity alone. Real-world evidence on the incremental benefit of adding GLP-1 RAs to background statin therapy in ASCVD is limited.

### What Is New?

In 119,911 propensity-matched ASCVD patients with elevated BMI on statins, GLP-1 RA use was associated with a 52% relative mortality reduction, 24% lower MI risk, and 34% lower stroke risk over 3 years. These associations were consistent in a BMI ≥35 subgroup and across four washout sensitivity analyses. A familial hypercholesterolemia exclusion sensitivity analysis preserved the mortality and stroke benefits and amplified the LDL <55 target achievement finding, supporting robustness of the primary results.

### What Is Next?

Prospective randomized trials evaluating GLP-1 RA addition to guideline-directed statin therapy as a formal combination regimen for ASCVD with obesity are needed. Studies with granular statin adherence and dose data should investigate the LDL >190 mg/dL signal. Mechanistic studies should evaluate whether GLP-1 RAs and statins exert additive anti-inflammatory and plaque-stabilizing effects.

## Data Availability

All data referred to in the manuscript were obtained from the TriNetX Research Network and are available from the corresponding author.

## REFERENCES

1. World Health Organization. Global Health Estimates: Life expectancy and leading causes of death and disability. Geneva, World Health Organization; 2020.

2. Jones JE, Tang KS, Barseghian A, Wong ND. Evolution of More Aggressive LDL-Cholesterol Targets and Therapies for Cardiovascular Disease Prevention. J Clin Med. 2023;12(23):7432. Published 2023 Nov 30. doi:10.3390/jcm12237432

3. Emmerich SD, Fryar CD, Stierman B, Ogden CL. Obesity and severe obesity prevalence in adults: United States, August 2021–August 2023. NCHS Data Brief, No. 508. Hyattsville, MD: National Center for Health Statistics; 2024. 10.15620/cdc/159281

4. Powell-Wiley TM, Poirier P, Burke LE, et al.Obesity and Cardiovascular Disease: A Scientific Statement From the American Heart Association. Circulation. 2021;143(21):e984–e1010. doi:10.1161/CIR.0000000000000973

5. Marso SP, Daniels GH, Brown-Frandsen K, et al.Liraglutide and cardiovascular outcomes in type 2 diabetes. N Engl J Med. 2016;375(4):311–322. 10.1056/NEJMoa1603827

6. Marso SP, Bain SC, Consoli A, et al. Semaglutide and cardiovascular outcomes in patients with type 2 diabetes. N Engl J Med. 2016;375(19):1834–1844. 10.1056/NEJMoa1607141

7. Gerstein HC, Colhoun HM, Dagenais GR, et al. Dulaglutide and cardiovascular outcomes in type 2 diabetes (REWIND): a double-blind, randomised placebo-controlled trial. Lancet. 2019;394(10193):121–130. 10.1016/S0140-6736(19)31149-3

8. Lincoff AM, Brown-Frandsen K, Colhoun HM, et al. Semaglutide and cardiovascular outcomes in obesity without diabetes. N Engl J Med. 2023;389(24):2221–2232. 10.1056/NEJMoa2307563

9. Sattar N, Lee MMY, Kristensen SL, et al. Cardiovascular, mortality, and kidney outcomes with GLP-1 receptor agonists in patients with type 2 diabetes: a systematic review and meta-analysis of randomised trials. Lancet Diabetes Endocrinol. 2021;9(10):653–662. 10.1016/S2213-8587(21)00203-5

10. Blumenthal RS, Morris PB, Gaudino M, et al. 2026 ACC/AHA/AACVPR/ABC/ACPM/ADA/AGS/APhA/ASPC/NLA/PCNA Guideline on the Management of Dyslipidemia: A Report of the American College of Cardiology/American Heart Association Joint Committee on Clinical Practice Guidelines. J Am Coll Cardiol. Published online March 13, 2026. doi:10.1016/j.jacc.2025.11.016

11. TriNetX. TriNetX Research Network platform: Compare Outcomes Analysis. Available at: https://trinetx.com. Accessed April 3, 2026.

12. Ridker PM, Danielson E, Fonseca FA, et al. Rosuvastatin to prevent vascular events in men and women with elevated C-reactive protein. N Engl J Med. 2008;359(21):2195–2207. doi:10.1056/NEJMoa0807646

13. Drucker DJ. The cardiovascular biology of glucagon-like peptide-1. Cell Metab. 2016;24(1):15–30. 10.1016/j.cmet.2016.06.009

14. Ma X, Liu Z, Ilyas I, et al. GLP-1 receptor agonists (GLP-1RAs): cardiovascular actions and therapeutic potential. Int J Biol Sci. 2021;17(8):2050–2068. 10.7150/ijbs.59965

15. Bu T, Sun Z, Pan Y, Deng X, Yuan G. Glucagon-Like Peptide-1: New Regulator in Lipid Metabolism. Diabetes Metab J. 2024;48(3):354–372. 10.4093/dmj.2023.0277

16. Phinney SD, Tang AB, Waggoner CR, Tezanos-Pinto RG, Davis PA. The transient hypercholesterolemia of major weight loss. Am J Clin Nutr. 1991;53(6):1404–1410. 10.1093/ajcn/53.6.1404

17. Suissa S. Immortal time bias in observational studies of drug effects. Pharmacoepidemiol Drug Saf. 2007;16(3):241–249. 10.1002/pds.1357

18. Ahmad Z, Agarwala A, Cuchel M, et al. Update on familial hypercholesterolemia: An expert clinical consensus from the National Lipid Association. J Clin Lipidol. 2026;20(4):708–737. doi:10.1016/j.jacl.2026.01.011

